# Understanding the Preferences of Young Women in Self-Sampling Interventions for STI Diagnosis: A Discrete Choice Experiment Protocol

**DOI:** 10.1101/2024.01.05.23299719

**Authors:** Ziningi N. Jaya, Witness Mapanga, Tivani P Mashamba-Thompson

**Affiliations:** School of Health Systems and Public Health, Faculty of Health Sciences, University of Pretoria, Pretoria, South Africa; Department of Biomedical Science, Faculty of Natural Science, Mangosuthu University of Technology, KwaZulu-Natal, South Africa; Faculty of Health Sciences, University of Pretoria, Pretoria, South Africa

**Keywords:** Sexually transmitted infections, underserved communities, self-sampling intervention, discrete choice experiment, user preferences

## Abstract

**Introduction:** Sexually transmitted infections (STIs) are a significant public health concern globally, particularly affecting young women. Early diagnosis and treatment are essential to reducing or stopping the continuous spread of infections and the development of the associated complications. Syndromic management, which is commonly used for STIs, presents several barriers, particularly for young women. This protocol is for a study that aims to understand young women’s preferences for a self-sampling intervention for STI diagnosis by using a Discrete Choice Experiment (DCE).

**Methods and analysis:** The following attributes of a self-sampling intervention were identified through a Nominal Group Technique: accessibility, education, confidentiality, self-sampling method, youth-friendliness, and cost. A pilot study involving 20 participants was conducted to refine the DCE questionnaire. A total of 196 young women from underserved communities will be recruited. The participants will be sampled from communities, stratified by settlement type and socioeconomic status. Data will be analysed using the multinomial logit model and mixed logit model to assess preferences and heterogeneity.

**Ethics and dissemination:** The study findings have the potential to inform policies for STI treatment and management to align healthcare services with user preferences. This can improve STI healthcare access for young women in underserved communities. Ethical approval was obtained, and results will be disseminated through peer-reviewed journals and health conferences.

**Strengths and limitations of this study:**

- DCEs provide a platform for users or consumers to express their preference for particular goods or services based on their attribute selection.
- Previously STI healthcare service provision has not been aligned with the preferences of young women. Therefore, this will reveal their preferences for a self-sampling intervention for STI healthcare and management.
- In instances where user preferences do not align with current practices for STI healthcare, this will provide an opportunity for policies to be reviewed and amended accordingly.
- This type of impact on STI healthcare aligns with goal 3.1 of South Africa’s National Strategic Plan which seeks to improve access to healthcare services for STIs and other diseases (1). It also aligns with goal three of the United Nations which seeks to improve access to healthcare for all and thus achieve universal healthcare coverage (2, 3).
- Since our study will be conducted on young women residing in underserved urban populations, our findings may not be a true reflection of young women from diverse communities.

## Introduction

Sexually transmitted infections (STIs) are a major public health problem in South Africa, particularly among young women, who constitute a large portion of the overall infections (4, 5, 6). Early diagnosis and treatment of STIs is crucial to prevent the spread of these infections and long-term complications which include sexual and reproductive health complications (7, 8, 9, 10). Although STI healthcare services are available at local healthcare facilities, individuals in resource-limited settings and underserved communities have limited access to quality basic services including healthcare (11, 12). Additionally, young women may be reluctant to access STI healthcare services in these communities due to various factors potentially related to the syndromic management of STIs.

Although widely used, particularly in low-and-middle-income countries (LMICs) syndromic management presents several challenges that impact STI healthcare seeking behaviour, particularly in young women (13). These factors include the inability to detect asymptomatic infection, failure to identify symptoms of STI, fear of being judged for being sexually active, fear of stigmatization, and discomfort with invasive associated genital examinations (13, 14). Self-sampling interventions have been proposed as a potential solution to eliminate challenges presented by syndromic management and increase access to STI screening services for young women in underserved communities (15, 16). The effectiveness and acceptability of self-sampling interventions are well understood. However, the preferred delivery method of self-sampling interventions based on user preferences has not been developed particularly in the South African context.

Discrete choice experiments (DCEs) are a method that is used to uncover people’s preferences for products, services or certain scenarios (17). It is an attributes-centred approach with a significant outcome of being able to quantify individuals’ trade-offs between attributes. Ultimately, DCEs uncover how much an individual is willing to forgo to gain more of another attribute (18, 19, 20). DCEs have been used in public health to understand and inform various significant healthcare-related decisions. For example in the United Kingdom, a DCE was used to assess patient preferences for attributes of primary care services which included appointment waiting time and provider continuity (21). This DCE helped to inform service design and resource allocation. In another study, a DCE was used to investigate the healthcare professional preferences for the allocation of resources in healthcare settings (22). The findings of this study guided the optimisation of resource allocation for decision-makers.

When considering the proven usefulness of self-sampling interventions as a tool to address challenges with access and screening of asymptomatic STIs, it is imperative to investigate user preferences for the delivery method. As such, the objective of this study is to develop a user-friendly self-sampling intervention for diagnosing STIs in young South African women from underserved communities using a DCE. A DCE involving young women aged 18-25 years from underserved communities in eThekwini District Municipality in KwaZulu-Natal, South Africa, will be utilised. It is anticipated that the findings of this study will contribute to the development of a user-friendly self-sampling intervention for STI screening that is tailored to the needs and preferences of young women from underserved communities in eThekwini District Municipality, KwaZulu-Natal, South Africa. This study is important because it addresses a critical gap in the literature on STI screening interventions in South Africa. Furthermore, it has the potential to contribute to the development of an effective and acceptable solution to increase access to STI screening services for young women in underserved communities.

### Aim

The main aim of this study is to utilise a DCE to determine young women’s most preferred self-sampling intervention for STI diagnosis. We particularly explore trade-offs between ease of accessibility and convenience, cost, education and normalisation, confidentiality and communication, self-sampling collection method, and youth-friendliness. To our knowledge, this is the first study to utilise a DCE to determine young women’s self-sampling preferences for STI diagnosis.

## Methods and analysis

### Identifying and defining attributes

Determining key attributes and levels for the DCE is an important step. Employing qualitative methods such as the nominal group technique (NGT) to select and frame attributes improves the significance and pertinence of the study findings (23). The number of key attributes must be kept at a reasonable number to avoid confusing participating individuals (24, 25). For simplicity, the number of attributes is maintained between four to eight (16).

### Nominal group technique

The key attributes for the self-sampling intervention were developed using two nominal group technique (NGT) co-creation workshops which were conducted on separate occasions. The NGT is a qualitative exploratory method combining the generation of ideas with the concept of enquiry within a small group (23, 24) often comprising six to twelve participants (25). Participants in one NGT comprised eight healthcare personnel involved in STI healthcare service provision at a primary healthcare clinic (PHC) located in underserved urban communities in eThekwini District Municipality. Another NGT comprised eight sexually active young women aged 18 -25 years residing in underserved urban communities in eThekwini District Municipality. In both NGTs, the participants were asked to identify barriers that hindered young women from accessing STI healthcare services. The identified barriers were then ranked from high priority to low priority according to the choice of each person. Once this was complete, NGT participants developed attributes for a self-sampling intervention that would address some of the barriers which were highlighted.

### One on one interviews

Following the NGT co-creation workshops, ten young women were interviewed to confirm the validity of the attributes identified during the NGT. The young women interviewed were aged 18 -25 years residing in underserved communities. The interviews did not yield any new information that contradicted what was already identified during the NGTs.

### Determining the list of attributes and preference levels

Ultimately a total of eight attributes emerged from the NGTs namely accessibility, education, communication, convenience, youth-friendliness, self-sampling method, and cost of self-sampling kit. An expert research panel was asked to review these attributes and they suggested a merging of a few which resulted in six attributes. The final list of attributes includes accessibility and convenience, education and normalisation, confidentiality and communication, self-sampling method, youth-friendliness, and cost of self-sampling kits. See Table 1 for a detailed list of attributes and their preference levels.

**Table 1:**
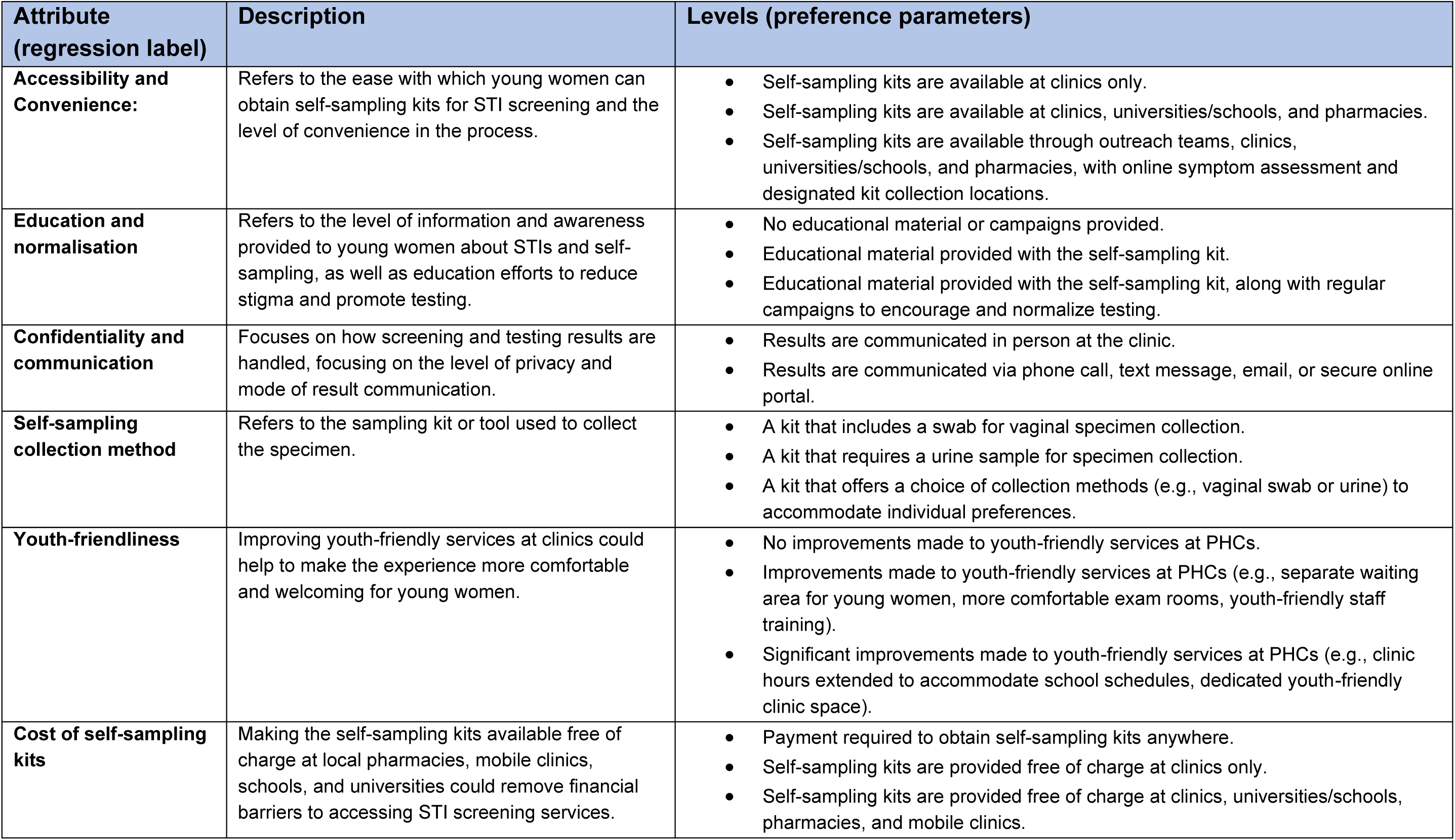
Attributes and levels.

| Attribute<br>(regression label) | Description | Levels (preference parameters) |
| --- | --- | --- |
| **Accessibility and Convenience:** | Refers to the ease with which young women can obtain self-sampling kits for STI screening and the level of convenience in the process. | • Self-sampling kits are available at clinics only.<br>• Self-sampling kits are available at clinics, universities/schools, and pharmacies.<br>• Self-sampling kits are available through outreach teams, clinics, universities/schools, and pharmacies, with online symptom assessment and designated kit collection locations. |
| **Education and normalisation** | Refers to the level of information and awareness provided to young women about STIs and self-sampling, as well as education efforts to reduce stigma and promote testing. | • No educational material or campaigns provided.<br>• Educational material provided with the self-sampling kit.<br>• Educational material provided with the self-sampling kit, along with regular campaigns to encourage and normalize testing. |
| **Confidentiality and communication** | Focuses on how screening and testing results are handled, focusing on the level of privacy and mode of result communication. | • Results are communicated in person at the clinic.<br>• Results are communicated via phone call, text message, email, or secure online portal. |
| **Self-sampling collection method** | Refers to the sampling kit or tool used to collect the specimen. | • A kit that includes a swab for vaginal specimen collection.<br>• A kit that requires a urine sample for specimen collection.<br>• A kit that offers a choice of collection methods (e.g., vaginal swab or urine) to accommodate individual preferences. |
| **Youth-friendliness** | Improving youth-friendly services at clinics could help to make the experience more comfortable and welcoming for young women. | • No improvements made to youth-friendly services at PHCs.<br>• Improvements made to youth-friendly services at PHCs (e.g., separate waiting area for young women, more comfortable exam rooms, youth-friendly staff training).<br>• Significant improvements made to youth-friendly services at PHCs (e.g., clinic hours extended to accommodate school schedules, dedicated youth-friendly clinic space). |
| **Cost of self-sampling kits** | Making the self-sampling kits available free of charge at local pharmacies, mobile clinics, schools, and universities could remove financial barriers to accessing STI screening services. | • Payment required to obtain self-sampling kits anywhere.<br>• Self-sampling kits are provided free of charge at clinics only.<br>• Self-sampling kits are provided free of charge at clinics, universities/schools, pharmacies, and mobile clinics. |

### Accessibility and convenience

Various studies report accessibility of healthcare services as a common challenge for young women (26, 27). By affording individuals the opportunity to self-collect specimens in a place that is convenient for them, self-sampling intervention improves accessibility (28, 29). Furthermore, in the age of technology, the use of online eHealth systems to improve access and convenience is well documented. As such, it was fitting for our NGT co-creation workshop participants to identify accessibility and convenience as an attribute for self-sampling interventions. We present the following choice or preference levels for self-sampling interventions to diagnose STIs in young women: making self-sampling kits available at clinics only; making self-sampling kits available at clinics, universities/schools, and pharmacies; or self-sampling kits available through outreach teams, clinics, universities/schools, and pharmacies, with online symptom assessment and designated kit collection locations.

### Education and normalisation

In the past health education campaigns have proved effective in destigmatising and normalising certain diseases as an intervention to encourage individuals to seek healthcare (30). Considering the stigma associated with STIs and barriers experienced by young people, health education campaigns have the potential to de-stigmatise and normalise these infections (31), and potentially improve healthcare seeking behaviour among this population. As an attribute of a self-sampling intervention, the main aim will be to educate the community about STIs and self-sampling as an intervention. We present the following choices or preference levels for this attribute: no educational material or campaigns provided; providing educational material together with the self-sampling kit; or providing educational material provided with the self-sampling kit, along with regular campaigns to encourage and normalize testing.

### Confidentiality and communication

The lack of confidentiality and invasion of privacy have previously been highlighted as barriers to young people accessing STI healthcare services (32, 33). To this effect self-sampling as an intervention provides privacy and autonomy and mitigates this barrier, and potentially improves STI healthcare seeking behaviour among young people (34). This attribute refers to being able to maintain confidentiality during the STI healthcare process from diagnosis, to communicating results and providing treatment and minimise interaction with healthcare personnel until the point of treatment where required. The following choice or preference levels are presented for this attribute: results are communicated in person at the clinic; or results are communicated via phone call, text message, email, or secure online portal.

### Self-sampling collection method

Since STIs are caused by various types of microorganisms including bacteria and viruses, an ideal specimen for diagnosis is one in which all these pathogens can be detected. Self-collected specimens that have been used for STI diagnosis include urine and vaginal swabs (35, 36). To accommodate the differing preferences, the following choice or preference parameters are recommended in the DCE: a kit that includes a swab for vaginal specimen collection; a kit that requires a urine sample for specimen collection; or a kit that offers a choice of collection methods (e.g., vaginal swab or urine) to accommodate individual preferences.

### Youth-friendliness

Previous studies have highlighted challenges related to the interaction of young people with healthcare workers at healthcare facilities, particularly with issues related to sexual and reproductive healthcare (37, 38). This has an impact on their healthcare seeking behaviour and as such negatively impacts healthcare outcomes. Improving youth-friendly services at clinics could help to make the experience more comfortable and welcoming for young women. As an attribute of self-sampling interventions, the following choice or preference levels are presented: no improvements made to youth-friendly services at clinics; improve youth-friendly services at clinics (e.g., separate waiting area for young women, more comfortable exam rooms, youth-friendly staff training); or significantly improve youth-friendly services (e.g., clinic hours extended to accommodate school schedules, have dedicated youth-friendly clinic space).

### Cost of self-sampling kits

Individuals in underserved communities are often faced with the plight of having limited access to basic resources. Therefore, there is a concern about the cost of self-sampling kits for the intervention, especially among underserved communities. Previous studies have reported on the feasibility of self-sampling interventions as an alternative to syndromic management (39), which may sometimes lead to overdiagnosis and overtreatment of patients. The current attribute is mindful of this and speaks of making the self-sampling kits available free of charge at locations that are easily accessible to young people. The following choice or preference levels are presented for this attribute: self-sampling kits are provided free of charge at clinics only; or self-sampling kits are provided free of charge at clinics, universities/ schools, pharmacies, and mobile clinics.

### Pilot study

#### Experimental design and development of choice tasks

A pilot study was conducted to pre-test the list of attributes and levels as identified by stakeholders. Although there is no clear consensus about the required number of choice sets for a DCE, the usual number is said to be between 8 and 16 (40, 41). Through a group consensus, the development of the choice tasks using the 6 attributes and choice set levels was done. The pilot survey consisted of 16 choice tasks based on the six attributes identified by our stakeholders during the NGT co-creation workshops. Since there were no known findings about young women’s preferences, null priors were assumed. Each choice task comprised a scenario for the participants to respond to with a choice set of their preference. See Box 1 below for an example of a choice task with the scenario and Table 2 is an example of a choice task:

**Table 2:**
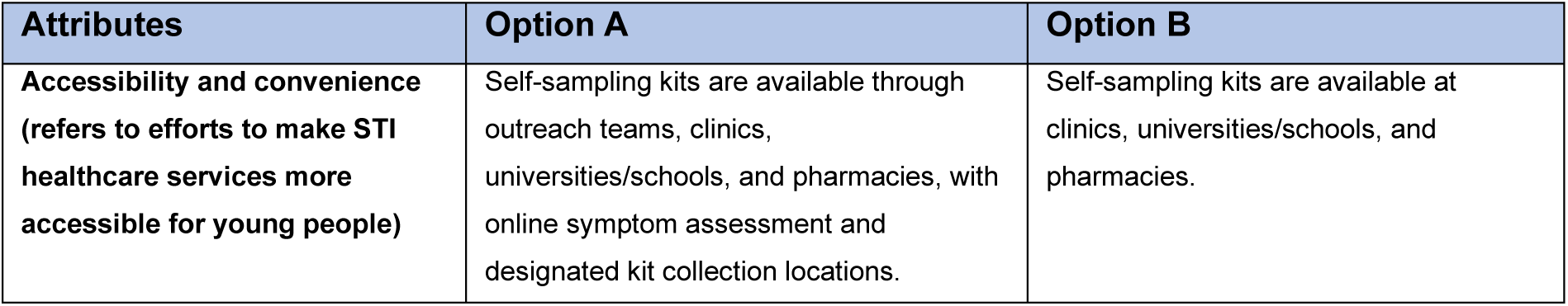
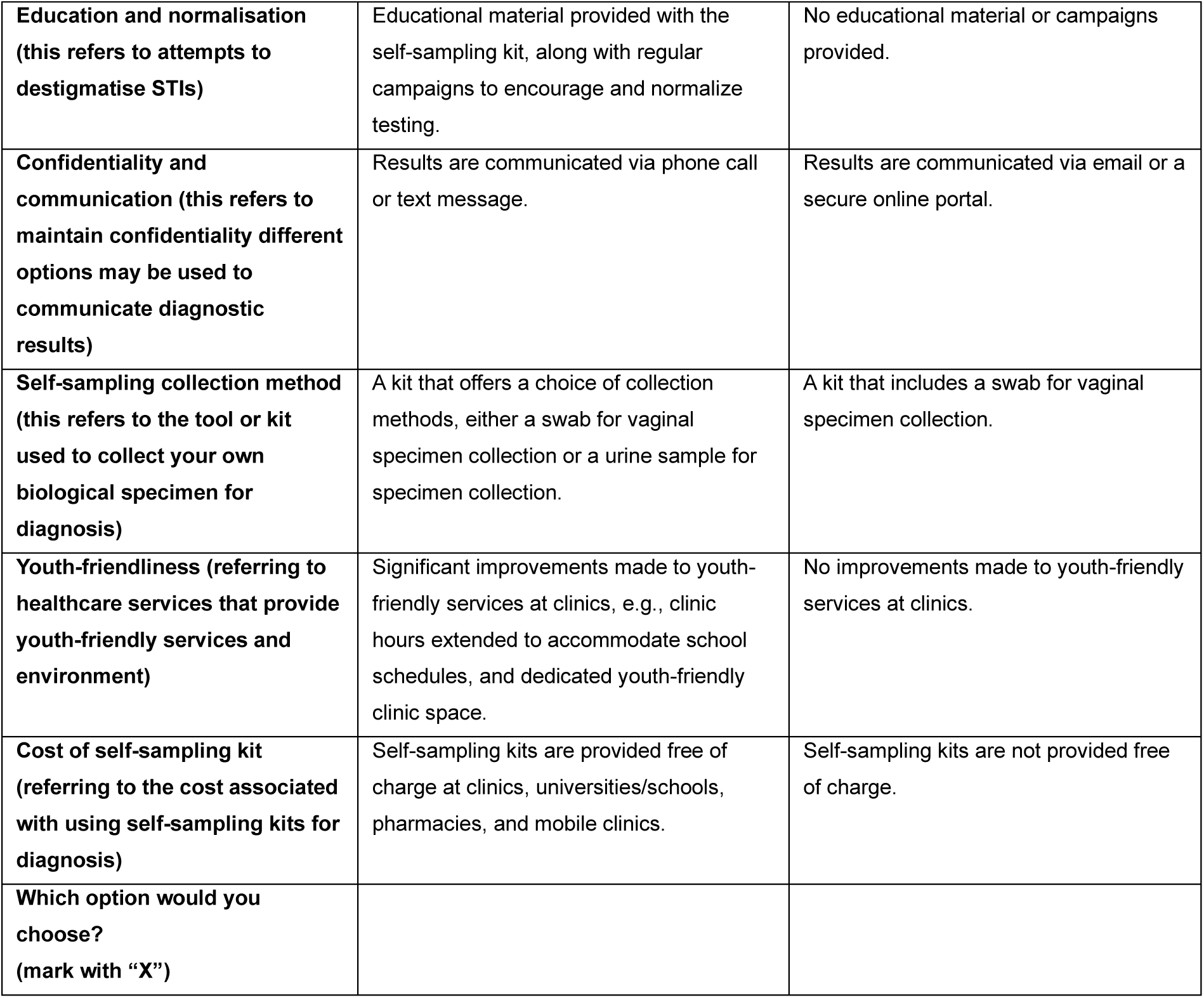
Example of a choice task.

| Attributes | Option A | Option B |
| --- | --- | --- |
| **Accessibility and convenience (refers to efforts to make STI healthcare services more accessible for young people)** | Self-sampling kits are available through outreach teams, clinics, universities/schools, and pharmacies, with online symptom assessment and designated kit collection locations. | Self-sampling kits are available at clinics, universities/schools, and pharmacies. |
| **Education and normalisation (this refers to attempts to destigmatise STIs)** | Educational material provided with the self-sampling kit, along with regular campaigns to encourage and normalize testing. | No educational material or campaigns provided. |
| **Confidentiality and communication (this refers to maintain confidentiality different options may be used to communicate diagnostic results)** | Results are communicated via phone call or text message. | Results are communicated via email or a secure online portal. |
| **Self-sampling collection method (this refers to the tool or kit used to collect your own biological specimen for diagnosis)** | A kit that offers a choice of collection methods, either a swab for vaginal specimen collection or a urine sample for specimen collection. | A kit that includes a swab for vaginal specimen collection. |
| **Youth-friendliness (referring to healthcare services that provide youth-friendly services and environment)** | Significant improvements made to youth-friendly services at clinics, e.g., clinic hours extended to accommodate school schedules, and dedicated youth-friendly clinic space. | No improvements made to youth-friendly services at clinics. |
| **Cost of self-sampling kit (referring to the cost associated with using self-sampling kits for diagnosis)** | Self-sampling kits are provided free of charge at clinics, universities/schools, pharmacies, and mobile clinics. | Self-sampling kits are not provided free of charge. |
| **Which option would you choose? (mark with "X")** |  |  |

##### Box 1

**Scenario for choice task**

###### Choice task scenario to contextualise the DCE

Imagine you are a young woman living in an underserved community, and you are considering getting tested for sexually transmitted infections. Self-sampling is a potential option for STI healthcare provision that allows you to collect your own specimen for laboratory diagnosis. It is an alternative current STI healthcare service that is fully facilitated by healthcare personnel in primary healthcare clinics. You are presented with options for a self-sampling intervention which include accessibility and convenience, education and normalisation, confidentiality and communication, self-sampling method, youth-friendliness, and cost of the self-sampling kit. Please consider the following choice task and select the option that is most suitable for you.

#### Pilot testing

Since there is no clear guidance on the sample size for DCE pilot studies, we utilised guidance by Bekker-Grob et al (42) which suggests that twenty to forty participants are sufficient for a pilot study. To satisfy our study, the pilot survey was distributed to thirty-five randomly selected young women aged 18 – 24 years residing in underserved communities in eThekwini District Municipality. Twenty young women completed the survey. Since this number is within the recommended total of twenty to forty participants, the pilot study data was accepted and analysed. The pilot tool was also used to determine the ease with which participants could complete the survey in terms of comprehension, and time taken to complete it. All participants reported ease and no comprehension challenges. However, 80% of participants reported that the tool was too long with a lot of choice tasks. They suggested reducing the number of choice tasks from sixteen to ten. All participants agreed that the attributes were all relevant and so did not need to change. The tool was amended accordingly based on participant comments.

#### Sampling and recruitment

Young women from underserved urban communities will be recruited for this study. Participant recruitment will be based on stratified random sampling where the underserved communities will be stratified into three subpopulations namely – core informal settlement, fringe informal settlement, and core township (43). The three strata will be defined according to the Council for Scientific and Industrial Research (CSIR) settlement typology of 2002 (43) as follows: core informal settlement refers to previously or currently illegal and unplanned settlements within inner cities or towns close to the traditional CBD or areas of employment, mostly with shacks as the predominant housing type; fringe informal settlement defined as freestanding, previously or currently illegal and unplanned settlements (mostly with shacks) located far away from the traditional CBD and often far from places of employment as well, resulting in extensive commuting patterns; and core township defined as Formal mass-built settlements (old or new) within inner cities or towns close to the traditional CBD or areas of employment. Furthermore, participant recruitment will also be based on socio-economic classification of households within the strata, and young women from poor households will be randomly selected.

The rule of thumb calculation as proposed by Johnson and Orme (44, 45) will be used to calculate the sample size for the experiment. The formula for the minimum sample size N calculation is as follows:

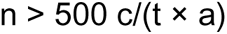

In the above equation, *c* is the largest number of levels for any one attribute; *t* represents the number of choice tasks; and *a* represents the number of alternatives in each choice task (45). Therefore, for our DCE using six attributes, with a maximum of three levels, and ten choice sets with two alternatives for each task, our required sample size is 75. Considering the wide range of data quality issues that have been reported for DCEs (46), we anticipate the exclusion of 30% of the respondents (47). As such we will increase our sample size by 30% to accommodate any data quality issues which increases our sample size to 98. We will investigate the heterogeneity of preferences and so we will double our sample size to 196 participants.

### Data analysis

Trade-offs between the attributes will be determined using the multinomial logit (MNL) model. By analysing participant preferences, it will help us to identify which factors influence participant preferences. The overall optimisation model will be optimised with the use of the MNL model as a framework (48). Although it is useful, the MNL model ignores heterogeneity and cannot manage random differences in individual preferences. However, the mixed logit model compensates for this shortfall because it does allow explanatory variables that are random (49). The mixed logit model will be used to investigate preferences between participants in the different strata. Presentation of results will include tables displaying coefficients for attribute levels and covariates, accompanied by pertinent statistical indicators such as pseudo R-squared, log likelihood test, and Akaike information criterion to assess model fit. Furthermore, the calculation of marginal rates of substitution, derived from the negative ratio between estimated coefficients, will provide insight into the relative importance of different attributes. This analysis will enable policymakers and clinicians to comprehend respondents’ willingness to trade-off certain attributes for the acquisition of others.

### Ethics and dissemination

Ethical approval was obtained from the University of Pretoria Research Ethics Committee (reference number 136:2022) and the KwaZulu-Natal Department of Health (reference number KZ_202208_005) before data collection. Written informed consent was obtained from all research participants who participated in the NGT. All participants who completed the pilot survey provided written consent prior to their participation. Written informed consent will be obtained from all participants prior to data collection for the main study. Research findings will be submitted to a peer-reviewed journal for publication. The research findings will also be presented at a relevant health conference.

## Acknowledgements

We would like to thank the healthcare workers and young women who participated in the NGTs which enabled the identification of attributes to be used in this DCE. We also thank the young women who participated in the pilot study.

## Authors’ contributions

Conceptualization, Z.N.J. and T.M.-T.; data collection; writing - original draft, Z.N.J.; writing—reviewing and editing, T.M.-T., and W.M; supervision, T.M.-T and W.M.

## Funding statement

This research received no specific grant from any funding agency in the public, commercial or not-for-profit sectors.

## Competing interests statement

None

## Patient consent for publication

obtained.

## Ethics approval

ethical clearance was obtained from the University of Pretoria Research Ethics Committee (reference number 136:2022) and the KwaZulu-Natal Department of Health (reference number KZ_202208_005) prior to data collection.

## Data availability statement

no additional data available.

## Notes

### Competing Interest Statement

The authors have declared no competing interest.

### Funding Statement

This study did not receive any funding

### Author Declarations

Ethics committee of the Faculty of Health Sciences (University of Pretoria) gave ethical approval for this work.

